# Inequalities in local funding cuts to environmental and regulatory service expenditure in England from 2009 to 2020

**DOI:** 10.1101/2024.01.26.24301656

**Authors:** Lauren Murrell, Katie Fahy, Helen. E Clough, Roger Gibb, Xingna Zhang, Marie Chattaway, Mark. A Green, Iain Buchan, Benjamin Barr, Daniel Hungerford

## Abstract

**Background:** Local authorities have been subject to substantial reductions in funding, placing strain on local services. Environmental and Regulatory (ER) services provide essential functions including infectious disease prevention and control via Food Safety and Animal and Public Health Infectious Disease Control services (APHIDC). This study investigates inequalities in local funding cuts to these services.

**Methods:** We used a generalised estimating equation model to estimate the annual percent change of ER service expenditure between 2009/10 and 2020/21 in addition to Food Safety and APHIDC expenditure change overall, and as a share of total ER expenditure. Models analysed trends by deprivation level, local authority structure and population density.

**Results:** Areas of higher deprivation had the largest reduction in expenditure, with ER and Food Safety and APHIDC cuts of 2% and of 23% respectively, compared to a 1% and 8% reduction in the least deprived areas. The share of ER expenditure spent on Food Safety and APHIDC decreased by 13% in the most deprived authorities compared to 6% in the least deprived areas. Environmental and Regulatory services saw the largest cuts in unitary authorities, declining by 2%. London boroughs had the greatest reductions in Food Safety and APHIDC expenditure, decreasing by 10%. Both ER and Food Safety and APHIDC expenditure decreased with increasing population density.

**Conclusion:** The unequal distribution of cuts shows the need for increased and equitable investment into these services to enable resilience to emerging infectious disease threats, and to prevent widening of health inequalities.

**Key Messages:** *What is already known on this topic:*

- Austerity measures have led to substantial reductions in local funding placing increased pressure on local authority services in England, research shows reductions vary by deprivation level of an area, rural - urban classification and local authority structure.
- It is unknown how local funding cuts to Environmental and Regulatory services, which provide essential services for public health protection, vary by these characteristics.

*What this study adds:*

- We investigate inequalities in austerity-enforced changes in Environmental and Regulatory service expenditure and sub-spending lines of Food Safety and Animal and Public Health Infectious Disease Control over time by deprivation, local authority structure and population density from 2009/10 to 2020/21.
- The largest cuts were in the more deprived areas and with increased population density for both Environmental and Regulatory and Food safety and Animal and Public Health Infectious Disease Control services. The largest cuts in Environmental and Regulatory services were seen in unitary authorities whereas Food safety and Animal and Public Health Infectious Disease Control saw largest cuts in London boroughs.

*How this study might affect research, practice or policy:*

- This research provides strong evidence of inequalities in local authority service expenditure in Environmental and Regulatory services and highlights where investment should be focused, in order to protect environmental and public health.

## Background

Over the past 10 years the UK has faced substantial changes to public spending at the local authority level. Following the financial crisis of 2008, austerity measures were implemented to reduce fiscal deficit and stabilise the economy (1). These measures led to significant cuts to local authority funding, which fell 77% between 2009-10 and 2020-21 (2,3). Local authorities are responsible for providing key services such as social services, housing, leisure and cultural services, and Environmental and Regulatory (ER) services (sometimes referred to as environmental health or regulatory services) (4). A growing body of literature describes the considerable impact of local funding cuts, demonstrating disproportionately grave impacts on disadvantaged areas (5–7), including increased disease burden (5).

Established in 1848, the UK Public Health Act empowers local authorities to protect the health of the population, addressing issues like infectious diseases, water safety and waste management (8). Environmental and Regulatory services align with the mandates of the Public Health Act, delivering water safety, waste collection, trading standards, food safety, and animal and infectious disease control (9). These services allow investment in the safeguarding of local public health via different routes, with ER services responsible for the notification and prevention of infectious disease (10). In 2020, ER services dealt with 58,434 notifiable incidents of infectious disease, with 4,800 cases due to food poisoning (8). Food safety services within ER include enforcement, inspection, regulation and monitoring of food premises and establishments (11), and investigation of food borne disease outbreaks and illness (9). Meanwhile, service expenditure under Animal and Public Health Infectious Disease Control (APHIDC) is focused on infectious disease control functions under the Public Health Act of 1984 (9).

Local authority funding comes from a combination of central government grants, local council tax, income from fees and charges. The distribution of these funds to services is decided by local government and differs by their structure and responsibilities. Unitary authorities, including London boroughs, are single-tier authorities and are responsible for all functions. Two-tier authorities split responsibilities between the upper (county) and lower (district) levels. Single-tier authorities can redirect funding from service budgets in order to protect statutory services such as Social Care. However, two-tier authorities are limited in their ability to redirect funding from services lower tier services, such as ER services, to upper tier services such as Social Care. Environmental and regulatory services have received substantial cuts since the introduction of austerity measures (2,12,13). These spending cuts have been accompanied by service changes, including reports of reductions in Food Safety staff by 13% between 2012/13 and 2017/18(14) a decline in food standards and hygiene sampling, and decreased waste removal (14–16). Existing research indicates that even before austerity, ER services were insufficient to meet the needs of the most deprived neighbourhoods (17,18). Furthermore, environmental health teams reported their capacity to function during the COVID-19 pandemic was impacted by prior local funding cuts (19). Therefore, expenditure reductions may further exacerbate inequalities in service provision and health outcomes.

The study aims to describe trends in ER service expenditure since the introduction of austerity, and how these have differed by level of neighbourhood socioeconomic deprivation, local authority structure and urbanicity service need.

## Methods

The study protocol containing further detail of study design, the analytical framework and data sources, and indicators have been previously published (20). We present a concise summary of the methods below.

### Setting

We analysed annual expenditure data in England from 2009/10 to 2020/21 for lower tier and single-tier local authorities. The geographical boundaries used in this analysis are based on 2020 local authority boundaries. The City of London and Isles of Scilly were excluded from the study due to their small populations and local characteristics. Additionally, for the analysis of Food Safety and APHIDC a further nine local authorities were removed due to missing data, leaving 303 for analysis.

### Data and measures of interest

The indicators of interest were total ER service expenditure, and the sub-streams of Food Safety and APHIDC expenditure. Data are provided at the lower tier local authority level each year from 2009/10 to 2020/21. These data were obtained from the Place Based Longitudinal Data Resource (PLDR) (21). Gross expenditure is defined as the total spending by local authorities, including income raised in service provision, and data are indicative of the financial year-April 1^st^ to March 31^st^. The midyear population estimate for each local authority was obtained from the Office of National Statistics (ONS) to estimate per capita expenditure.

English Indices of multiple deprivation (IMD) were used as the measure of deprivation (22). Population density estimates, obtained from ONS (24), were originally measured in persons per square kilometre. Population density was divided by 1000, so that a unit increase corresponded to a change of 1000 persons per square kilometre. This facilitates interpretation of model estimates as a unit of one person per square kilometre is less meaningful.

Population density may influence infectious disease risk due to greater transmission in urban areas. Additionally, it acts as an indicator of food outlet density. Direct data on food outlet density was poor quality with inconsistent reporting; in addition, there was a significant correlation between population density and the number of food establishments at the local authority level (Pearson Correlation Coefficient 0.76, p<0.0001). Therefore, population density was used as a more robust proxy indicator for service need.

### Analysis

Expenditure data were adjusted for inflation prior to analysis using gross domestic product (GDP) deflator (23). Local authorities were designated to deprivation quintiles using IMD 2019 and held consistent temporally, enabling comparability, as deprivation levels in some local authorities have changed over time. The local authority IMD average score was used to produce quintiles weighted by population size, making the number of people in each quintile evenly distributed. Local authorities were also grouped into three categories, London boroughs, lower tier districts or unitary (including metropolitan) authorities, allowing comparison across local authority structure.

Data were analysed at the aggregated ER service level, then explored by individual spending line. The sub-streams of Food Safety and APHIDC were then explored, investigating how expenditure changed over time by deprivation, population density (classed as rural or urban), and by local authority structure in exploratory analysis.

To measure temporal trends in expenditure, data were stratified by deprivation, local authority structure and population density. A generalised estimating equation (GEE), with a log link function (24) was fitted to assess temporal trends in expenditure and our explanatory variables (24). The outcome was annual gross total ER expenditure, with log-transformed population size as an offset, in order to model the log of per capita spend. Financial year was specified to interact with local authority structure, population density and deprivation level. This model was then reproduced for gross Food Safety and APHIDC service expenditure. The proportion of Food Safety and APHIDC expenditure as a share of total gross ER expenditure was explored, with Food Safety and APHIDC as the outcome variable and log transformed gross ER expenditure as the offset. We interpreted the GEE model results by calculating the linear combinations of the estimated coefficients. The results were presented as annual percentage change.

### Missing data and multiple imputation

Missing data were present in the Food Safety and APHIDC services for 17 local authorities. Of these, nine were removed if data were missing for three consecutive years, two consecutive years at the beginning of the study period (2009 and 2010), or at the end of the study period (2019 and 2020) as specified in the logic model (online supplementary figure S4). Eight were then carried forward for multiple imputation with 10 observations missing, using predictive mean matching, and a predictor matrix consisting of deprivation level, local authority type, and population data to estimate missing values. Sensitivity analysis was carried out with case wise deletion of all authorities with missing values (online supplementary table S3).

## Results

### Trends in expenditure

There was a decline in mean expenditure for ER and Food Safety and APHIDC per capita between 2009/10 and 2020/21, across each deprivation level, local authority type and rural and urban local authorities (table 1). The most deprived local authorities, London boroughs and urban authorities saw the largest reductions in mean expenditure per capita for ER and Food Safety and APHIDC expenditure.

**Table 1.** Mean expenditure in 2009/10 and 2020/21 summary table.

|  | Baseline ER mean expenditure per capita 2009/10, £ (SD) | Change in mean ER expenditure per capita 2009/10-2020/21, £ (%) | Baseline Food Safety + Animal Infectious Disease Control expenditure per capita 2009/10, £ (SD) | Change in mean Food Safety + Animal Infectious Disease Control expenditure per capita 2009/10-2020/21, £ (%) | Number of Local authorities | Mean population density per square kilometre (SD) |
| --- | --- | --- | --- | --- | --- | --- |
|  | IMD quintile |  |  |  |  |  |
| **1 (Least deprived)** | 144.35 (21.52) | -15.02 (-10.4%) | 8.74 (6.04) | -5.09 (-58.2%) | 88 | 581(820) |
| **2** | 155.84 (21.08) | -22.74 (-14.6%) | 8.41 (5.08) | -4.39 (-52.2%) | 67 | 1338(1729) |
| **3** | 169.24 (60.57) | -24.49 (-14.5%) | 8.37 (5.39) | -5.28 (-63.2%) | 68 | 2035(2802) |
| **4** | 176.23 (47.17) | -37.85 (-21.5%) | 10.84 (8.57) | -7.32 (-67.5%) | 45 | 3429(3779) |
| **5 (Most deprived)** | 179.16 (35.2) | -48.64 (-27.1%) | 9.37 (6.59) | -6.67 (-71.2%) | 44 | 2551(2435) |
|  | Local authority structure |  |  |  |  |  |
| **London Borough** | 217.82 (83.88) | -50.37 (-23.1%) | 10.75 (7.21) | -7.75 (-72.1%) | 32 | 7344(3760) |
| **Two tier** | 153.38 (23.92) | -17.26 (-11.3%) | 9.2(6.24) | -5.24 (-57.0%) | 188 | 769(923) |
| **Unitary (excluding London)** | 159.34 (30.15) | -38.00 (-23.8%) | 7.92 (5.78) | -5.33 (-67.35%) | 92 | 1793(1434) |
|  | Rural or Urban |  |  |  |  |  |
| **Rural** | 153.18 (23.52) | -13.71 (-9%) | 9.35 (5.61) | -5.42 (-58%) | 92 | 152(61) |
| **Urban** | 165.56 (46.23) | -32.47 (-19.6%) | 8.82 (6.52) | -5.56 (-63%) | 220 | 2428(2745) |

### Environmental and regulatory service expenditure

There is variation in the direction and size of expenditure change between local authorities. Most local authorities experienced an overall decline in expenditure, with the largest being 57%. However, 33 Local authorities experienced an increase in ER expenditure in 2020/21 compared to 2009/10, with the largest being 60% (figure 1).

**Figure 1.**
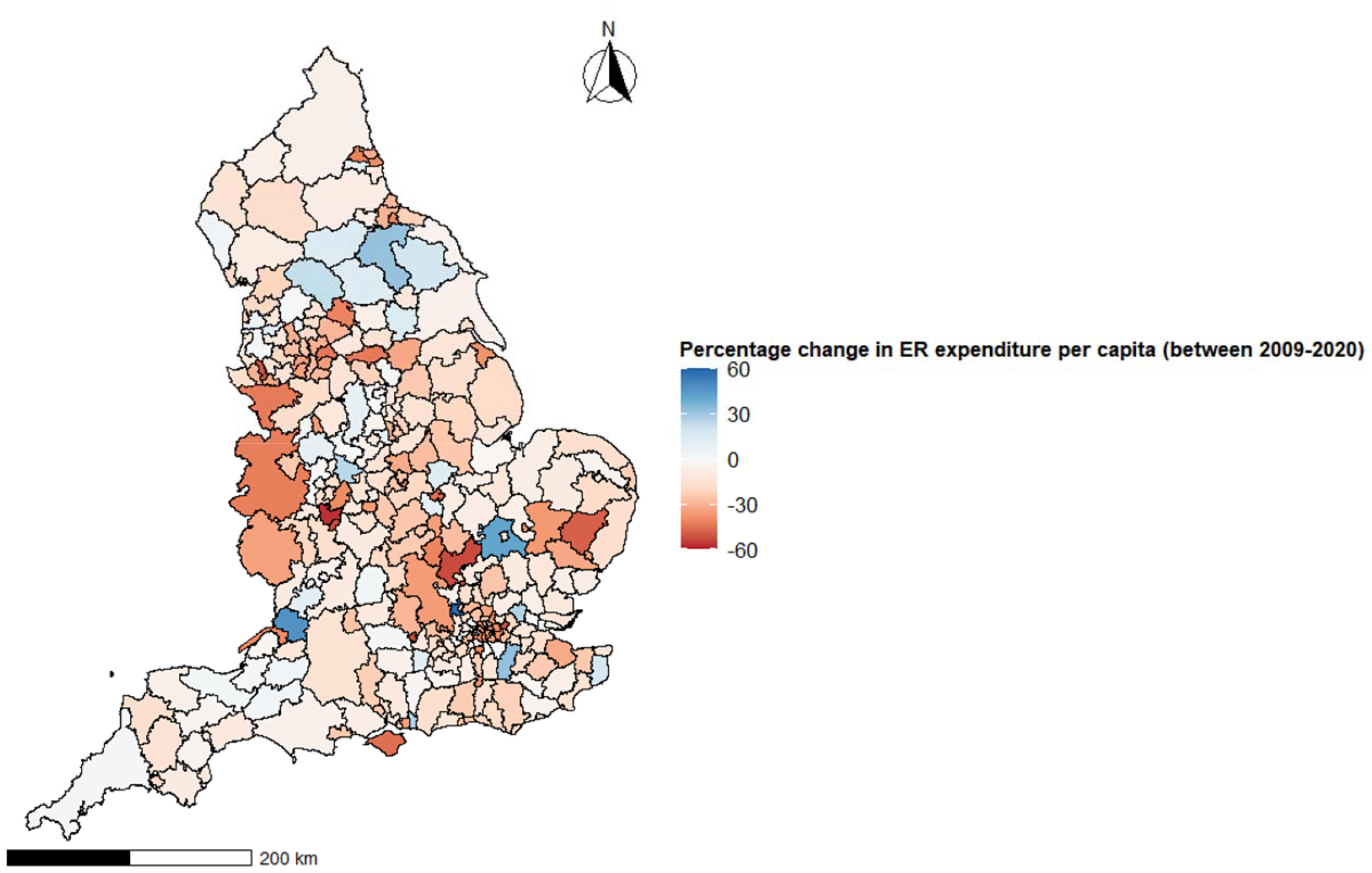
The percentage change in Environmental and Regulatory service expenditure between 2009 and 2020 at the local authority level.

The gross ER service expenditure per capita decreased by 27% in the most deprived local authorities, from 2009/10 to 2020/21, compared to an 10% decrease in the least deprived local authorities (figure 2). In London boroughs and unitary authorities, expenditure experienced a larger decline compared to two-tier local authorities, which had a maximum reduction of 10% in 2020/21. By 2012, London boroughs and unitary authorities surpassed this decline, both experiencing reductions of over 23%. During this period relative cuts to gross ER service expenditure per capita in urban areas were almost double that in rural areas.

**Figure 2.**
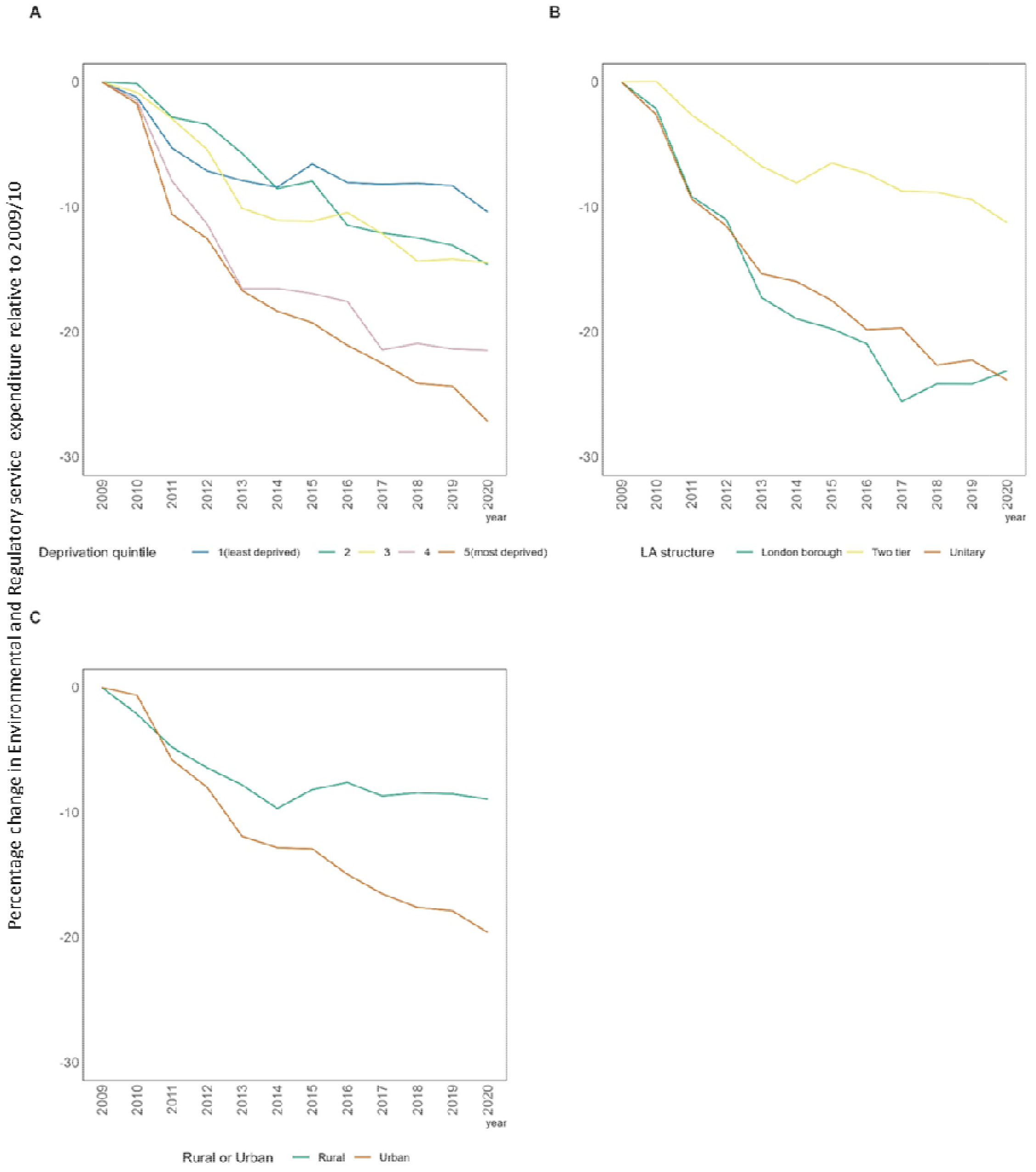
The percentage change in mean ER expenditure per capita relative to 2009/10, stratified by deprivation, local authority structure and rural urban.

### Food Safety and Animal Public Health and infectious disease control expenditure

There was a steep decline in expenditure on Food Safety and APHIDC services, with a decrease of 71% by 2020/21 in the most deprived local authorities (Figure 3). Compared to other local authority structures, London boroughs saw the largest cuts in expenditure overtime, with 2020/21 cut by 72 % relative to 2009/10. Two tier local authorities saw the smallest reduction in expenditure of 57% between 2009/10 and 2020/21. Rural and urban local authorities see a matched decline until 2012, from this point urban areas see a moderately larger decline in annual expenditure, decreasing by 63%, compared to 58% in rural areas.

**Figure 3.**
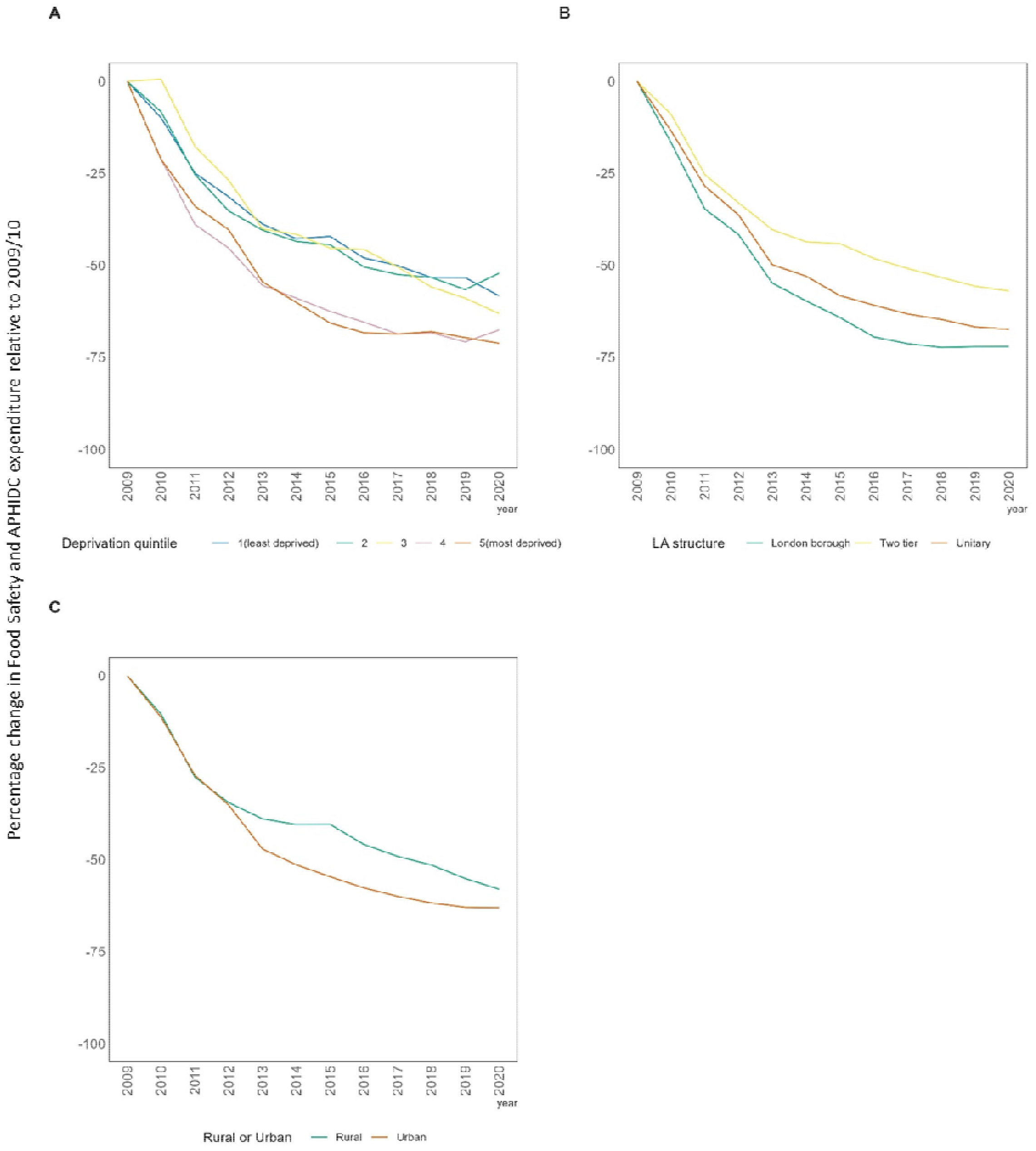
The percentage change in mean Food Safety and APHIDC expenditure per capita relative to 2009/10, stratified by deprivation, local authority structure and rural urban.

### Generalised estimating equation expenditure trends

The estimated trends in ER service expenditure per capita from the GEE models, show annual declines in expenditure per capita (table 2). The most deprived local authorities experienced a reduction of 2.4% (95% CI: -3.5%, -1.3%) in expenditure annually compared to 1.2% (95% CI: -1.8%, -0.5%) in the least deprived local authorities. Trends in expenditure also varied between local authority structure. The largest cuts were in unitary authorities, which saw an annual decrease of 1.9% (95% CI: -2.9%, -0.8%) in expenditure, compared to a decrease of 1% (95% CI: -3%, 1%) in London boroughs. For each additional 1000 people per square kilometre there was decrease of 1.4% (95% CI 95% CI: -2.1% -0.7%) in expenditure annually.

**Table 2:**
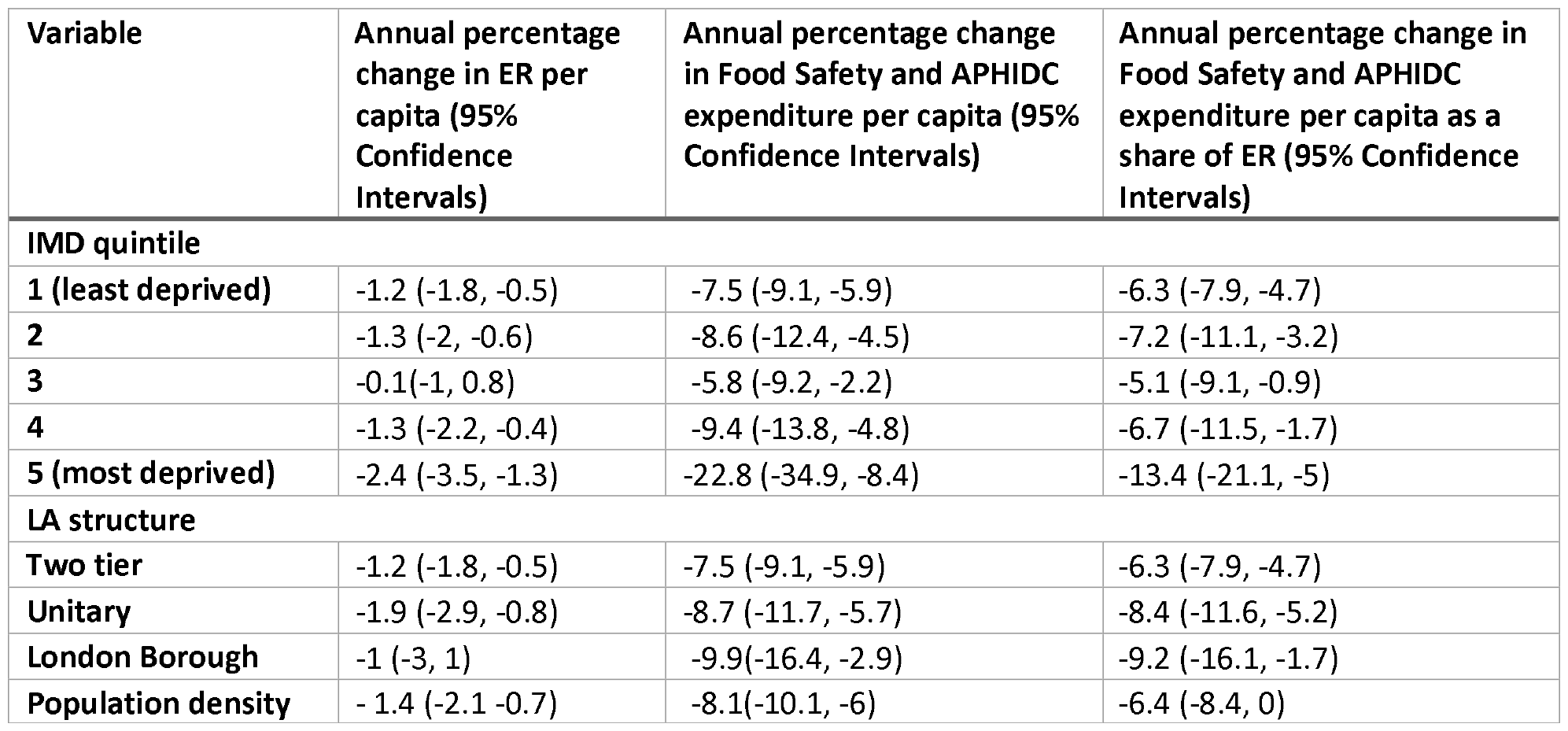
Estimates of annual percentage change in ER, Food Safety and APHIDC expenditure per capita from generalised estimating equation models, between 2009/10 and 2020/21.

The greatest decrease in Food Safety and APHIDC expenditure was in the most deprived local authorities (−22.8%, 95% CI: -34.9%, -8.4%) annually compared to the least deprived (−7.5%, 95% CI: -9.1%, -5.9%). Of local authority structures, London boroughs experienced the largest reductions with a decrease in expenditure per capita of 9.9% (95% CI: -16.4%, -2.9%). There was a decrease in Food Safety and APHIDC expenditure of 8.1% (95%CI: -10.1%, -6%) annually for each additional 1000 people per km^2^ increase in population density.

The share of expenditure spent on Food Safety and APHIDC decreased by 13.4% (95% CI: -21.1%, -5%) per capita annually in the most deprived authorities compared to 6.3 % (95% CI: -7.9%, -4.7% in the least deprived. The share of ER service expenditure per capita spent on Food Safety and APHIDC was cut the most in London boroughs (−9.2% 95% CI: -16.1%, -1.7%) and decreased by 6.4% (95% CI: -8.4%, 0%) per capita annually, per 1000 increase in population density.

## Discussion

### Summary

In this longitudinal study we investigate post-austerity inequalities ER service expenditure trends and spending lines of Food Safety and APHIDC. Our study demonstrates how austerity has exacerbated social and spatial inequalities in environmental public health, which could lead to widening health inequalities. Expenditure for total ER services decreased more in the most deprived areas, unitary local authorities, and local authorities with higher population density. These inequalities in expenditure cuts are accentuated when analysing Food Safety and APHIDC, with most deprived areas experiencing the largest reductions over time and as a share of total ER service expenditure. Expenditure for Food Safety and APHIDC decreased most in London boroughs and unitary authorities compared to two-tier authorities which saw the smallest reductions, overall and as a share of ER service expenditure. Furthermore, overall Food Safety and APHIDC expenditure decreased in areas with higher population density, as did the share of ER service spent on Food Safety and APHIDC.

### Interpretation

Reductions in expenditure across ER services and within Food Safety and APHIDC were largest in the most deprived local authorities compared to the least deprived, supporting previous findings on expenditure cuts (7,25). These reductions were larger for Food Safety and APHIDC services, annual decreases of 22.8% compared to 2.4% for overall ER. This suggests a variation between the sub-service streams, which might be missed in higher level analysis. A report by the National Audit Office describes differences in spending between ER sub-services, with community safety falling by 47.1% compared to 11% for water safety (15), highlighting the importance of sub-service level analysis. Statutory services such as those carried out by ER services have seen smaller reductions compared to other services (15,26). However, local authorities facing larger cuts are less able to protect these services from reductions (15). Therefore, with the increasing demand on social care (16), a protected service, areas with higher demands for this service may see smaller expenditure available for ER services, meaning more deprived areas could see larger cuts to statutory services such as ER. In addition, more deprived local authorities may struggle to generate income outside of central government funding (their primary source of funding), through means such council tax and business rates due to factors such as lower property values and lower business rates. So inequitable cuts to statutory service funding will result in inequitable service delivery and impact, with research showing that food establishments in the most deprived areas are 25% less likely to obtain required hygiene standards than those in the least deprived areas (28). Furthermore, individuals in more deprived areas are of greater risk of gastrointestinal infections (GI infection) (29), and experience more severe outcomes (30,31). This inequality in infection risk is particularly true for pathogens associated with person-to-person transmission and some foodborne pathogens (29). These trends may be of particular concern, as more deprived areas tend to have a higher demand for Food Safety and APHIDC services but have experienced the largest reduction in funding.

Our study also found the largest reductions in ER service expenditure to be in unitary authorities. Whilst London boroughs and unitary authorities saw the largest reductions in Food Safety and APHIDC expenditure, overall and as a share of ER service expenditure. This is in line with previous research by Fahy et al, 2022 which describes the largest reduction to Cultural, Environmental and Planning (CEP) in unitary authorities (25). This is likely due to single-tier authorities making larger cuts to and within the ER service budget to protect higher priority services, such as Social Care and exacerbated by the different capacities to subsidise services with alternative revenues (13). For instance, in 2018/19 unitary local authorities generated 15% of spending need from fees and charges compared to 60% in district councils (27). Furthermore, in unitary authorities, council tax contributes less to total income compared to county councils (17).

Findings showed that as population density increased, so did cuts in overall ER service expenditure, Food Safety and APHIDC, and the as a share of ER service expenditure. This is in line with previous research that showed urban areas in England have faced more substantial reductions in CEP spending compared to rural (25). Budgets in areas with higher population density may see larger cuts due to larger reliance on central government funding in urban areas compared to rural areas, which have alternative means of income generation (2). This is of concern as areas with higher population density are likely to have higher food establishments density, increasing the service need in these areas.

### Strengths and Limitations

This study is the first to spatially and temporally analyse expenditure of ER services using systematic statistical methods. We utilise national data for these services, including the analysis of critical sub-services, whilst accounting for population change and deflation, thus providing a comprehensive indication of trends in expenditure overtime. This provides evidence, which may be used in informing policy and direction of needs at a larger scale. Study limitations include uncertainty surrounding sub-service reporting accuracy and the missing data within these services. It is possible that misclassification and reporting errors occurred within reporting of local authority expenditure resulting in some sub-services reporting zero expenditure for a given year. Furthermore, the difference in missingness may be due to the discretionary characteristics of a service meaning they are not prioritised, and funding may be directed elsewhere. We tried to account for this by combining sub-services of interest, in addition to using multiple imputation. Another limitation is the use of population density as a proxy of food establishment density. However, high correlation between these data and well reported population density made it a suitable measure. Finally, this study is descriptive, therefore, we cannot confirm why these trends have occurred or what the implications of these findings might be, this should be investigated in future work.

## Conclusion

Despite the importance of ER services as a key component of local health protection systems, they have been cut substantially year-on-year since the introduction of austerity. The concerning trend that cuts to these protective services have hit more deprived, highly populated-urban areas, presents a high risk for resilience to current and future infectious disease threats. This observed decline is unsustainable and threatens the ability for this service to function, posing a potential threat to the public from infectious diseases. Further research is needed to understand the impact on health outcomes of these inequitable cuts to ER services. Additionally, increased, and equitable investment to these services is urgently needed to protect public health.

## Data Availability

The data sets used in for analysis are available at https://pldr.org/, https://www.ons.gov.uk/ and https://www.gov.uk/

https://www.gov.uk/

https://www.ons.gov.uk/

## Data availability

The expenditure data included in this study are available from https://pldr.org/ (21), further information and guidance for the data can be found here. Population demographic data can be found at https://www.ons.gov.uk/ and https://www.gov.uk/

## Acknowledgements

We gratefully acknowledge the contributions of Alexandros Alexiou, Frieda Jorgenson, Simon Melican, Nick Wellington, Samantha Walters, Valerie Decraene, Jane Muizelaar

## Ethics

Ethics approval is not applicable, data used in this study is open access available to the public.

## Contributions

LM, HEC, RG, XZ, MAG, MC, IB, BB, DH were involved in conceptualization of the study. LM and DH were involved in investigation, and project administration. LM, KF, DH, HEC, MAG, BB and XZ contributed to methodology. LM carried out visualisation, writing – original draft preparation. LM, KF, HEC, RG, XZ, MAG, MC, IB, BB, DH contributed to writing-reviewing and editing. HEC, RG, XZ, MAG, MC, IB, BB, DH contributed to supervision. BB was responsible for funding acquisition.

## Abbreviations

ER: Environmental and Regulatory Services
APHIDC: Animal and Public Health Infectious Disease Control
LA: Local Authority
NAO: National Audit Office
ONS: Office of National Statistics
IMD: Index of Multiple Deprivation
UK: United Kingdom
GEE: Generalised Estimating Equation
GI: Gastrointestinal
PLDR: Place Based Longitudinal Data Resource

