## Supplementary material for "Inequalities in local funding cuts to environmental and regulatory service expenditure in England from 2009 to 2020": Online Supplemental material

### 1. Definitions

Indices of deprivation use seven domains to provide measures of deprivation at the lower layer super output area; income deprivation, employment deprivation, education, skills and training deprivation, health deprivation and disability, crime, barriers to housing and services, living environment deprivation(1)

### 2. ER spending line categories.

- Food safety gross
- Water safety gross
- Trading standards gross
- Port health gross
- Public conveniences gross
- Animal and Public health infectious disease control gross
- Pest control gross
- Environmental protection noise and nuisance gross
- Defence agianst flooding
- Land drainage and related work
- Coas protection
- Agricultural and fisheries
- Street cleanin not chargeable to highways
- Waste collection
- Waste disposal
- Trade waste
- Recycling
- Waste minimisation
- Climate change
- Cemerty cremation and mortuary
- Housing standards
- Health and safety
- Liscening - alcohol and enterainment liscencing; taxi liscencing
- Crime reduction
- Safety services

### 3. Environmental and Regulatory expenditure

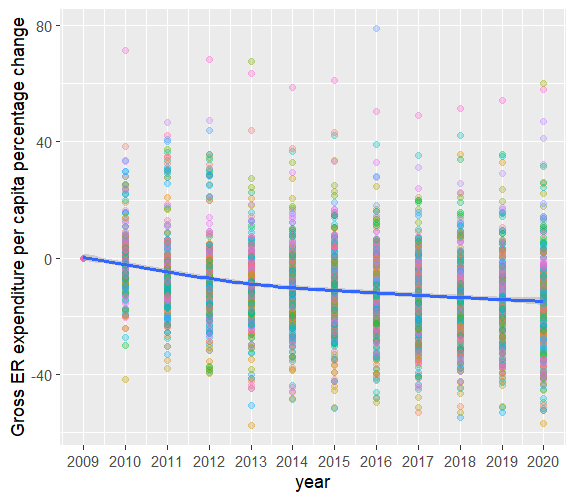

*Figure 1* Percent change in Environmental and Regulatory service expenditure annually by local authority, relative to 2009. Each circle corresponds to an individual local authority, blue line represents the average change each year.

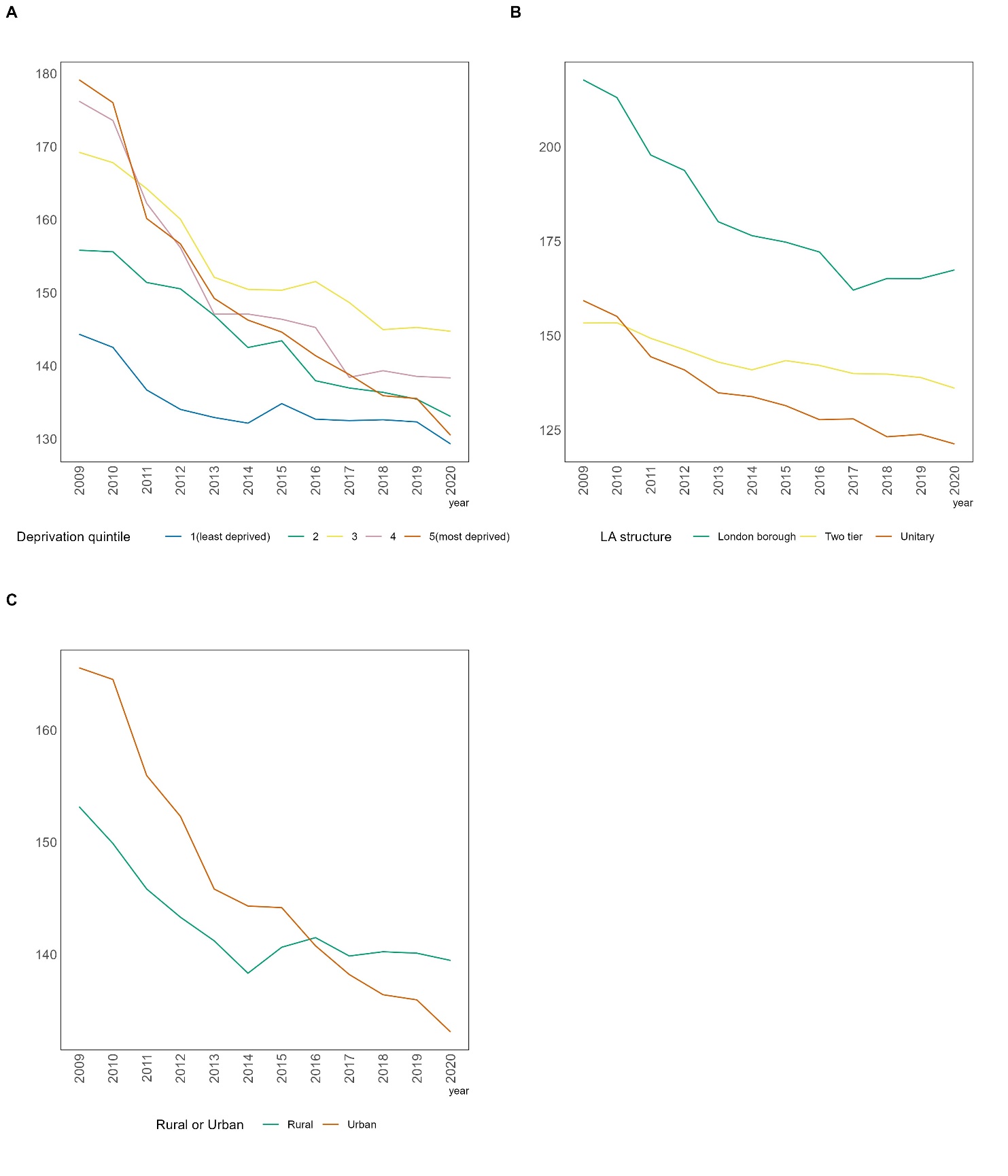

*Figure 2* Average Environmental and Regulatory expenditure (£) per capita between 2009 and 2020, stratified by deprivation quintile, local authority structure, rural and urban.

### Food Safety + APHIDC expenditure

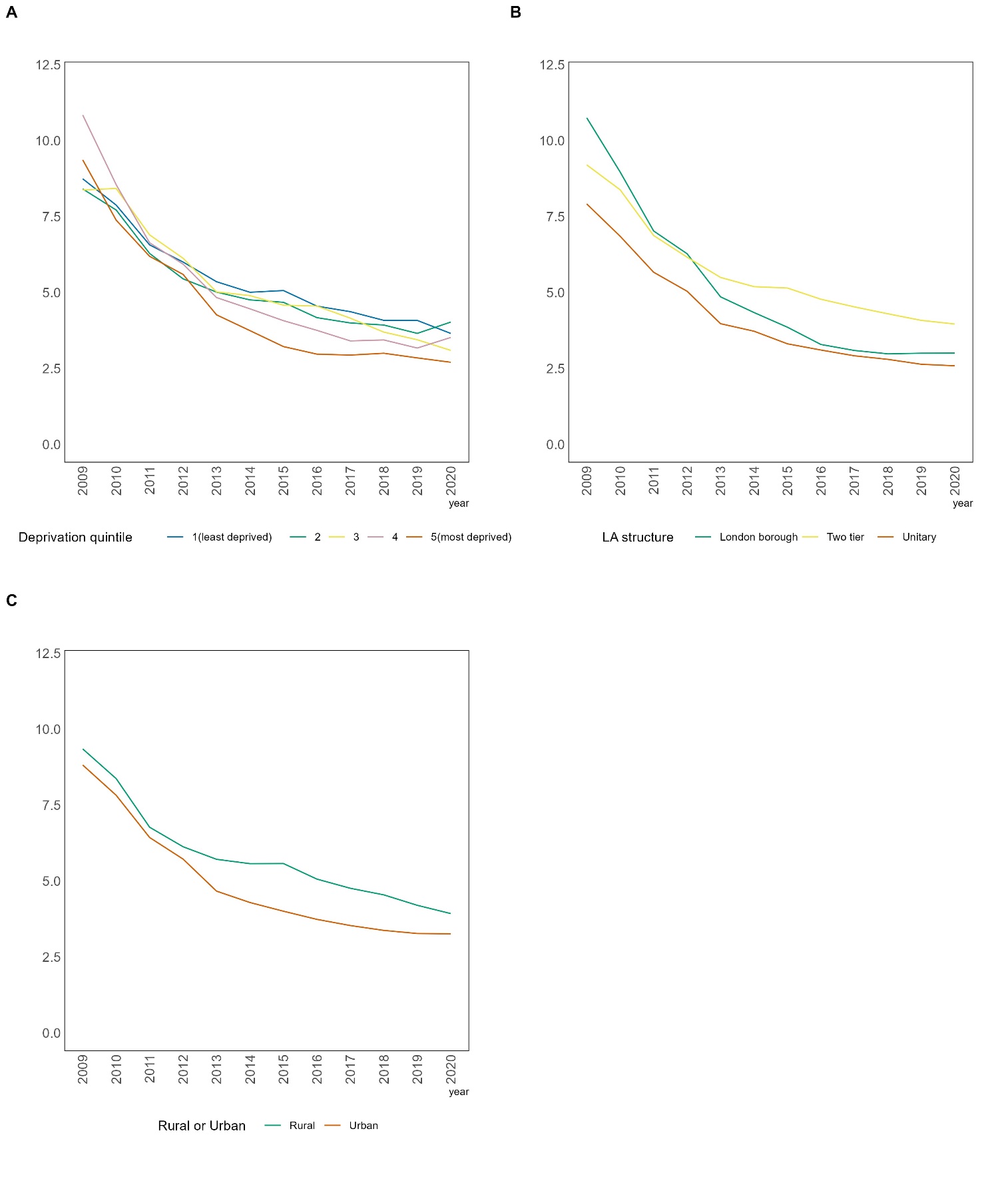

*Figure 3* Average Food Safety and APHIDC (£) per capita between 2009 and 2020, stratified by deprivation quintile, local authority structure, rural and urban.

### Missing data

##### Summary table of missing data within ER services

Table 1

| Environmental and Regulatory Sub-streams | % of observations reported as 0 |
| --- | --- |
| Agricultural and fisheries services gross | 54.87 |
| Animal and public health infectious disease control gross | 7.44 |
| Cctv gross | 29.91 |
| Cemetery cremation and mortuary services gross | 7.81 |
| Climate change costs gross | 59.71 |
| Coast protection gross | 76.62 |
| Crime reduction gross | 11.58 |
| Defences against flooding gross | 37.93 |
| Environmental protection noise and nuisance gross | 8.94 |
| Food safety and animal and public health and infectious disease control | 1.15 |
| Food safety gross | 6.26 |
| Health and safety gross | 28.92 |
| Housing standards gross | 54.68 |
| Land drainage and related work gross | 44.36 |
| Licensing alcohol and entertainment licensing taxi licensing gross | 3.24 |
| Pest control gross | 19.18 |
| Port health gross | 93.63 |
| Public conveniences gross | 10.73 |
| Recycling gross | 7.49 |
| Safety services gross | 29.91 |
| Street cleansing not chargeable to highways gross | 1.07 |
| Trade waste gross | 33.6 |
| Trading standards gross | 0.45 |
| Waste collection gross | 0.51 |
| Waste disposal gross | 0.05 |
| Waste minimisation gross | 37.32 |
| Water safety gross | 74 |

### Flow diagram for exclusion (Multiple Imputation)

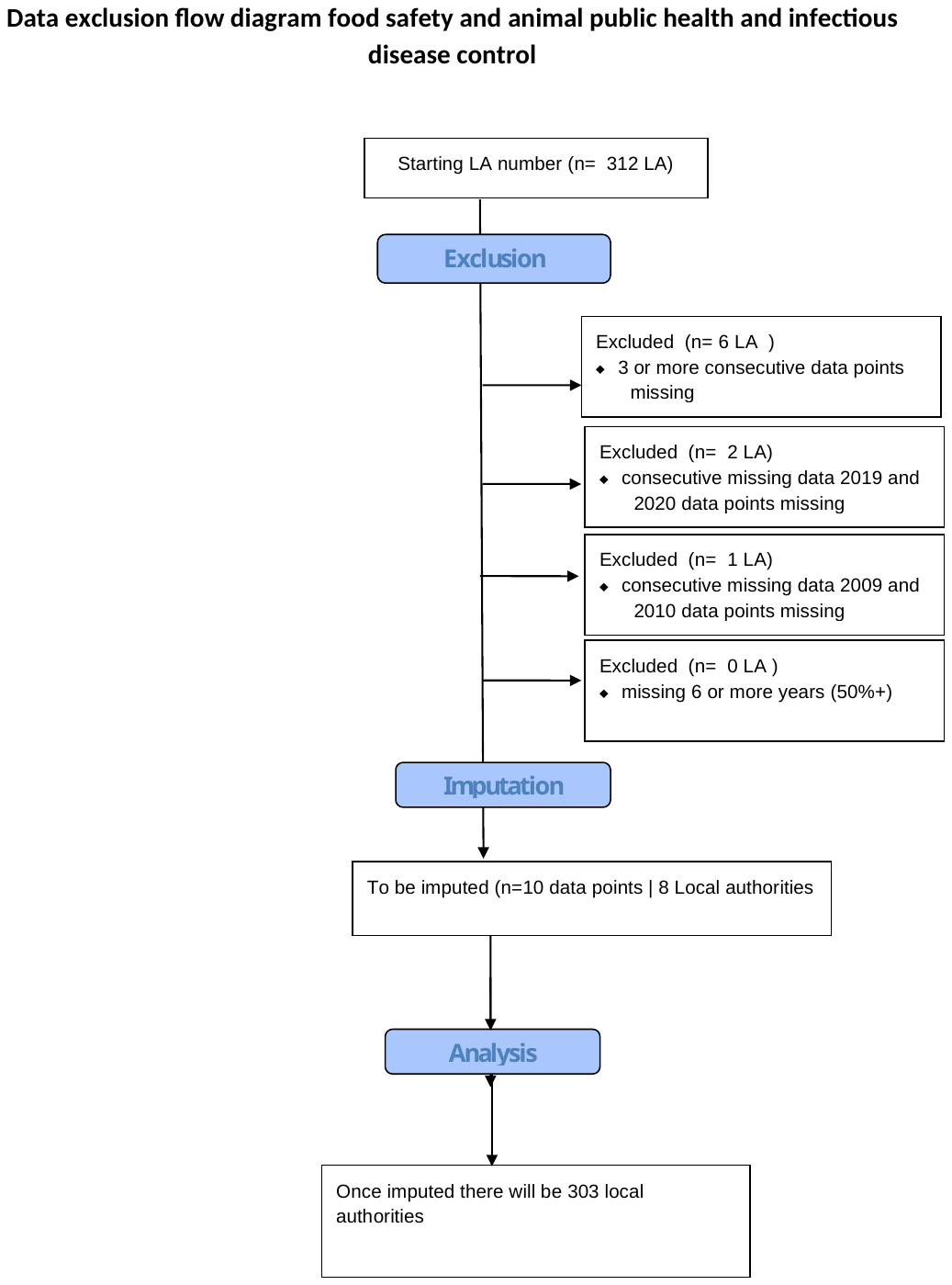

*Figure 4* Flow diagram displaying data exclusion logic model for excluding local authorities from statistical analysis.

Table 2: Local authorities removed or included for multiple imputation.

| Excluded Local authorities | | Local authorities carried forward for multiple imputation | |
| --- | --- | --- | --- |
| LA code | LA name | LA code | LA name |
| E06000016 | Leicester | E06000042 | Milton Keynes |
| E06000035 | the Medway towns | E07000086 | Eastleigh |
| E06000045 | Southampton | E07000111 | Sevenoaks |
| E06000056 | Central Bedfordshire | E07000116 | Tunbridge wells |
| E08000008 | Tameside | E07000166 | Richmond shire |
| E09000011 | Greenwich | E07000177 | Cherwell |
| E06000007 | Warrington | E09000008 | Croydon |
| E06000012 | Northeast Lincolnshire | E09000030 | Tower hamlets |
| E07000066 | Basildon |  |  |

##### Density plot 1:

Density plot 1 shows the observed values (blue), and estimated values from multiple imputation (red).

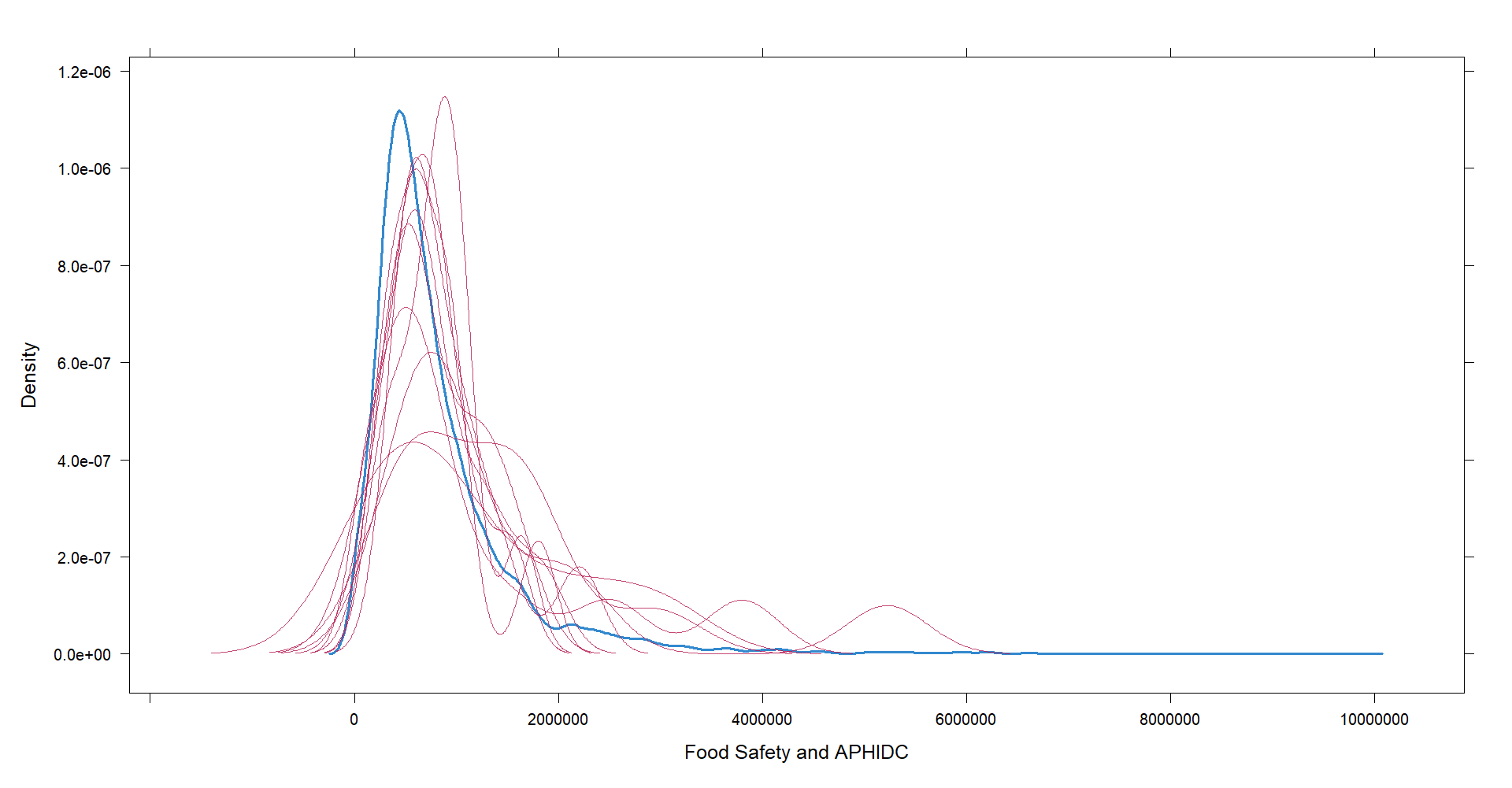

*Figure 5* Density plot showing observed values (blue) and estimated values (red) from multiple imputation.

### Sensitivity analysis

#### GEE:

###### Sensitivity analysis: GEE results for case wise deletion

Table 3: Estimates of annual percentage change in Food Safety and APHIDC expenditure per capita from generalised estimating equation models, between 2009/10 and 2020/21 using case wise deletion (removal of all local authorities with reports of 0 for any year/ service)

| Variable | Annual percentage change in Food Safety +APHIDC expenditure per capita (95% Confidence Intervals) | Annual percentage change in Food Safety + APHIDC expenditure per capita as a share of ER (95% Confidence Intervals) |
| --- | --- | --- |
| 1 (least deprived) | -7.7(-9.3, -6.1) | -6.4 (-8, -4.8) |
| 2 | -9.2, (-12.3, -5.9) | -7.9 (-11.1, -4.6) |
| 3 | -6.2 (-8.8, -3.6) | -4.6 (-7.1, -1.9) |
| 4 | -9.1(-13.3, -4.6) | -6.1( -10.6, -1.3) |
| 5 (most deprived) | -24.1 (-36, -10) | -13.7 (-21, -5.8) |
| Two tier | -7.8(-9.3, -6.2) | 6.4 (-8.0, -4.8) |
| Unitary | -9.3 (-11.6, -6.9) | -8.8 (-11.2, -6.4) |
| London Borough | -10.7(-17.1, -3.8) | -7.8 (-14, -1.1) |
| Population density | -7.8 (-9.5, -6.1) | -6.4 (-8.1, -4.6) |
